# Does HIV impact susceptibility to COVID-19 (SARS-CoV-2) infection and pathology? A review of the current literature

**DOI:** 10.1101/2020.12.04.20240218

**Authors:** Elnara Aghakishiyeva, Derek Macallan

## Abstract

**Objectives:** Giving appropriate guidance to people living with HIV (PLWH) during the COVID-19 pandemic depends on having adequate data to inform recommendations. Several studies have now been published which inform such advice. The objective of this study was to collate this information and review the implications of emerging data.

**Methods:** We performed a systematic literature search of studies relating COVID-19 to HIV infection from the beginning of the pandemic to end of November 2020. We included both published and pre-published manuscripts and analysed papers according to whether they primarily informed risk of infection or risk of adverse outcome.

**Results:** 68 papers (including 11 pre-prints) were identified. In terms of risk of infection, it appears that PLWH are no more or less likely to become infected with COVID-19. In terms of outcomes and mortality, most early small studies did not demonstrate an increase in mortality compared to background populations. However, several larger, more recent studies from South Africa, New York and two from the UK demonstrate higher mortality among PLWH when results are adjusted for other risk factors, giving relative risks of 2.1, 1.2, 1.7 and 2.3 respectively. Apparently conflicting results may arise from differences between studies in their power to account for cofactors and confounding variables. HIV-positive non-survivors tend to be younger and have fewer comorbidities than their HIV-negative counterparts; mortality may be higher in PLWH with low CD4 counts.

**Conclusions:** Although the literature appears conflicting, large studies which account for covariates strongly suggest that HIV infection increases COVID-19 mortality.

## Introduction

The ongoing coronavirus disease-2019 (COVID-19) pandemic, caused by severe acute respiratory syndrome coronavirus-2 (SARS-CoV-2), has emerged as a major global health threat. Two striking features of this disease are, firstly, its often high but variable infectivity and, secondly, the striking range in its clinical consequences. In terms of infectivity, even remote and pre-symptomatic contact may result in infection, (1) whereas highly-exposed household contacts may remain uninfected. Similarly, pathology is highly variable; some infected individuals remain asymptomatic, whereas others develop life-threatening multi-organ failure. Given that the virus is relatively stable, and that this spectrum seems largely independent of the immediate environment, such variability must derive predominantly from host factors. Several host susceptibility factors were recognised early in the pandemic and our understanding of risk factors for serious pathology has evolved rapidly over the last six months. We now recognise that the disease disproportionally affects older people, those with obesity and those with underlying health conditions such as diabetes, renal and cardiovascular disease. (2) This information has allowed the identification of highly-susceptible individuals and driven national and regional guidelines for “shielding” specific groups.

Whether HIV-infected individuals also fall into this “susceptible” group remains a matter for debate. This is a non-trivial issue as some people living with HIV (PLWH) have seen their freedoms curtailed “for their own good”, but guidance has varied widely between countries. Are such restrictions necessary or helpful? Early in the pandemic, in the absence of epidemiological evidence, advice followed theoretical reasoning: ‘PLWH have impaired immunity so are likely to be more susceptible to COVID-19’. However, in situations where most PLWH are taking antiretroviral treatment (ART) - which we know repletes lymphocyte subsets, restores clinical immunocompetence and endues normal life expectancy – a counter-argument might be made: ‘PWLH on effective treatment may be considered no more “susceptible” than the general population’. Some guidelines, distinguished between those on effective treatment and those not; this was the approach taken by BHIVA who advised a more cautious approach for those with CD4 counts <50 cells/Ul. (3) Basing advice on theoretical considerations is however fraught with difficulty in this setting. Firstly, PLWH are very heterogenous; secondly, even those on long-term ART still have a degree of immune dysfunction; thirdly, it might be argued that since most COVID-19 pathology is immunopathology, HIV infection may attenuate rather than exaggerate disease severity; finally, some anti-retroviral drugs taken by PLWH may have direct anti-viral effects on SARS-CoV2. Such theoretical complexity underlines the need for good observational data. In the absence of such data, many international guidelines followed a cautious approach in order to protect PLWH from COVID-related morbidity and mortality. (3,4)

The object of this paper is to summarise the currently available published or pre-published data on the susceptibility of PLWH to COVID-19. In order to evaluate published data, we utilise a model that separates the two risks that contribute to the probability of having an adverse clinical outcome from COVID-19 (Figure 1). If we denote ‘risk of exposure to COVID-19’ by *α*, ‘risk of becoming infected’ by *β, and ‘risk of adverse clinical outcome’ by *γ*, the probability of developing an adverse outcome from COVID-19 infection can be expressed as *α x β x γ* (Figure 1). Here α, β and γ are probability coefficients which range between 0 and 1. Shielding seeks to reduce α and is beyond the scope of this review. We will focus on evidence that informs us whether β and/or γ are higher for PLWH than for the general population. Firstly, we will consider whether* HIV increases susceptibility to SARS-CoV-2 infection; in terms of our model, ‘Is β_HIV_>β_0_?’ (using the suffix to denote the population, HIV being PLWH and 0 being the background population). Secondly, we will ask, does HIV increase the risk of COVID-19 infection developing into severe infection with adverse outcomes? In our model, ‘Is γ_HIV_>γ_0_?’ Thirdly, we will ask whether the literature supports risk stratification between PLWH according to CD4 counts and viral load. Such data are important because they provide the evidence base to inform national and regional guidance.

**Figure 1.**
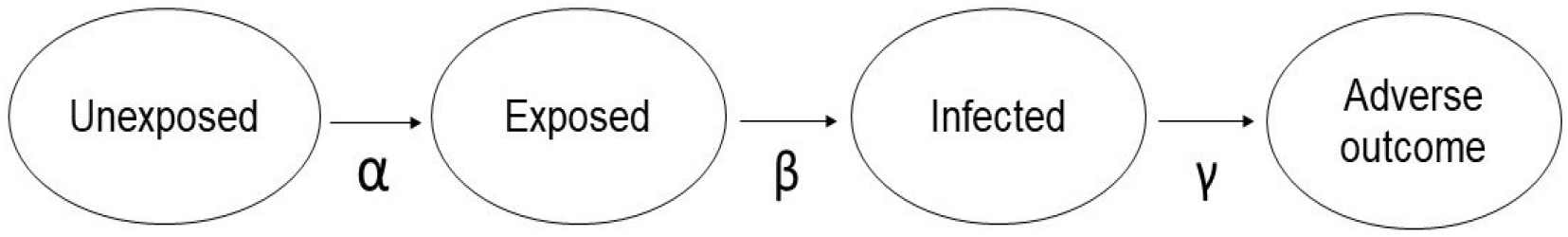
A model describing the probability of developing an adverse outcome from COVID-19. α, risk of exposure to COVID-19; β, risk of contracting COVID-19; γ, risk of developing a severe infection resulting in an adverse outcome.

## Methods

A review of relevant literature published between December 2019 and a final census date at the end of November 2020 was undertaken. We identified studies using keywords, ‘COVID-19’, ‘coronavirus disease-19’, ‘SARS-CoV-2’, ‘novel coronavirus’, ‘new coronavirus’, ‘severe acute respiratory syndrome coronavirus 2’, ‘people living with HIV’, ‘HIV/ AIDS’, ‘co-infection’, ‘immunosuppression’ in electronic databases: PubMed, Google Scholar and ScienceDirect, as well as direct searches in specific journals: The Lancet, JAMA, The BMJ, NEJM, Clinical Infectious Diseases, The Journal of Infectious Diseases. Since COVID-19 is a new disease, first identified in December 2019, some recent studies are not yet published. Therefore, the same search terms were used to search for literature on the preprint servers, Medrxiv and Biorxiv. For both published and pre-published studies the selection criteria included primary studies on HIV/ COVID-19 co-infection cases and secondary review studies. For the purposes of this analysis, we present only studies published in English. In total, we identified 68 studies, of which 54 were peer-reviewed, while 11 were preprints and 2 are conference abstracts.

## Results

### Does HIV increase the susceptibility to SARS-CoV-2 infection?

In order to answer this question, is β_HIV_>β_0_, we have to rely on the “natural experiment” of pandemic exposure. Since many SARS-CoV-2 infections are asymptomatic, one would ideally compare the prevalence of swab PCR-positivity in PLWH versus the background population regardless of symptoms, assuming similar levels of exposure and assuming a similar magnitude and duration of PCR-positivity (both of which may be untrue). To date, although many community prevalence studies have been performed, to our knowledge, none have yet reported prevalence or incidence of swab-test positivity against HIV status except where prompted by symptoms.

The next-best “natural experiment” is to see if PLWH are over-represented in patients being diagnosed with COVID-19? Of course, this is not completely independent of the severity question (is γ_HIV_>γ_0_?) since swabbing is usually prompted by the individual accessing healthcare, usually because of symptoms; this will introduce bias if thresholds for testing differ between PLWH and controls. Several studies have reported such data; a summary of HIV/ COVID-19 co-infection studies with prevalence data is shown in Table 1. Most are small and do not specifically focus on the relationship between HIV infection and COVID-19 susceptibility. Richardson *et al* published a study early in the epidemic that reported the clinical characteristics of 5,700 sequential patients hospitalized with COVID-19. (5) Of these, 43 were HIV seropositive, an HIV prevalence rate of 0.8%, similar to that in the community (∼0.9% from other sources).(6) More recent studies, conducted mainly in USA and Spain, report similar results. (7-11) These observations tally with our own experience (albeit biased by need for hospitalisation); of 1,584 admissions to our tertiary centre with COVID-19 during the first wave of the pandemic, 11 were seropositive (0.7%), a similar proportion to the HIV prevalence of our locality (0.54%).

**Table 1.**
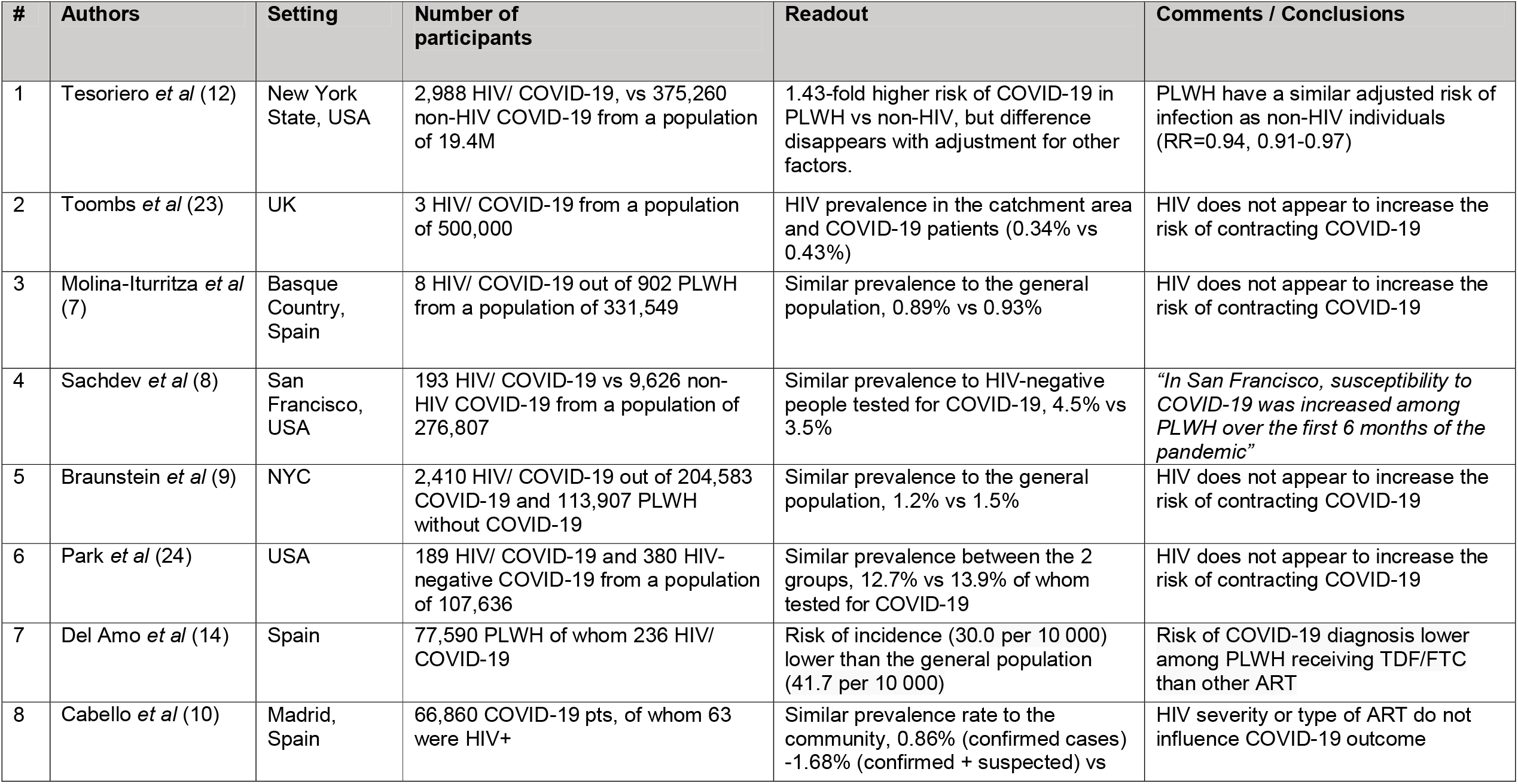
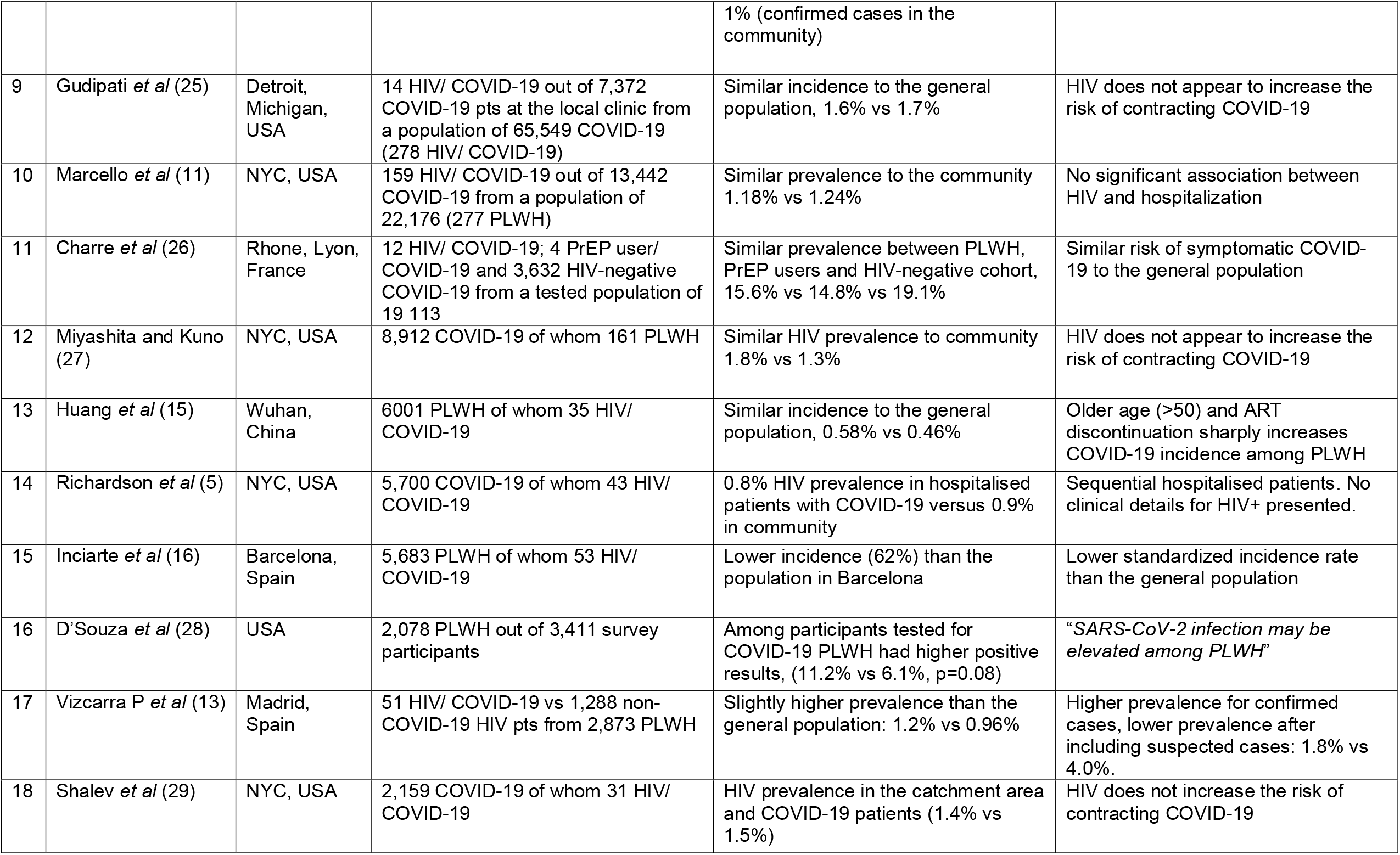
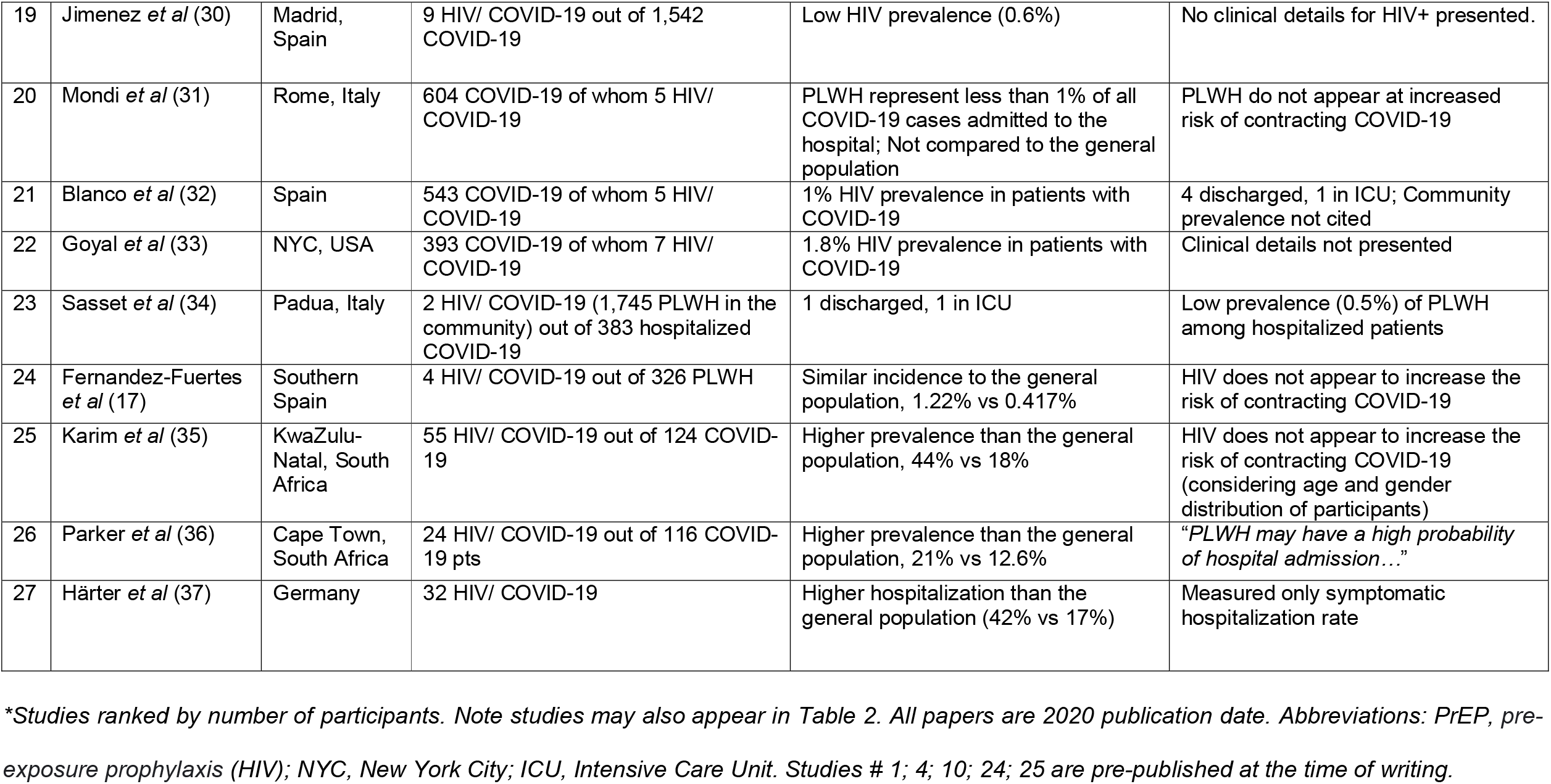
HIV/ COVID-19 co-infection studies with prevalence data.

A much larger, more-recent population-based study in New York state captured information on COVID diagnoses from across the state (over 19M people), so should not be biased by need for hospitalisation, but may be influenced by reason for swabbing. (12) Strikingly, although rates of COVID diagnosis were higher overall in PLWH (2.8% versus 1.9%), when adjusted for other risk factors (age, region of residence, race and ethnicity), the relative risk (RR) was virtually the same as non-HIV infected members of the same community (RR=0.94, 0.91-0.97). (12)

The converse approach is to investigate the incidence of COVID-19 in PLWH versus the general population (rather than the prevalence of HIV in COVID-19 incident cases as above). One Spanish study for example reported 51 co-infections from an HIV clinic population of 2,873 patients. (13) Including all suspected cases, gave a lower incidence rate in PLWH, 1.8% versus 4.0% in the community. However, rates were similar when only ‘confirmed case’ were considered (1.2% versus 0.9%), although confirmatory testing rates differed between PLWH (69%) and others (22%). Other studies have taken a similar approach reaching similar conclusions. (7, 14-17) Taken together, both prevalence (of HIV) and incidence (of COVID-19) studies seem to point to a rate of infection in PLWH similar to the background population.

### Does HIV increase the risk of severe infection with adverse outcomes in COVID-19 infection?

The second question we sought to address from current literature is whether, once infected, PLWH suffer more adverse outcomes than a comparable population without HIV. In our model this is represented by: ‘Is γ_HIV_>γ_0_’? Detailed clinical outcomes are difficult to define and compare; hence most studies focus on mortality data. A summary of HIV/ COVID-19 co-infection studies with mortality data is shown in Table 2.

**Table 2.**
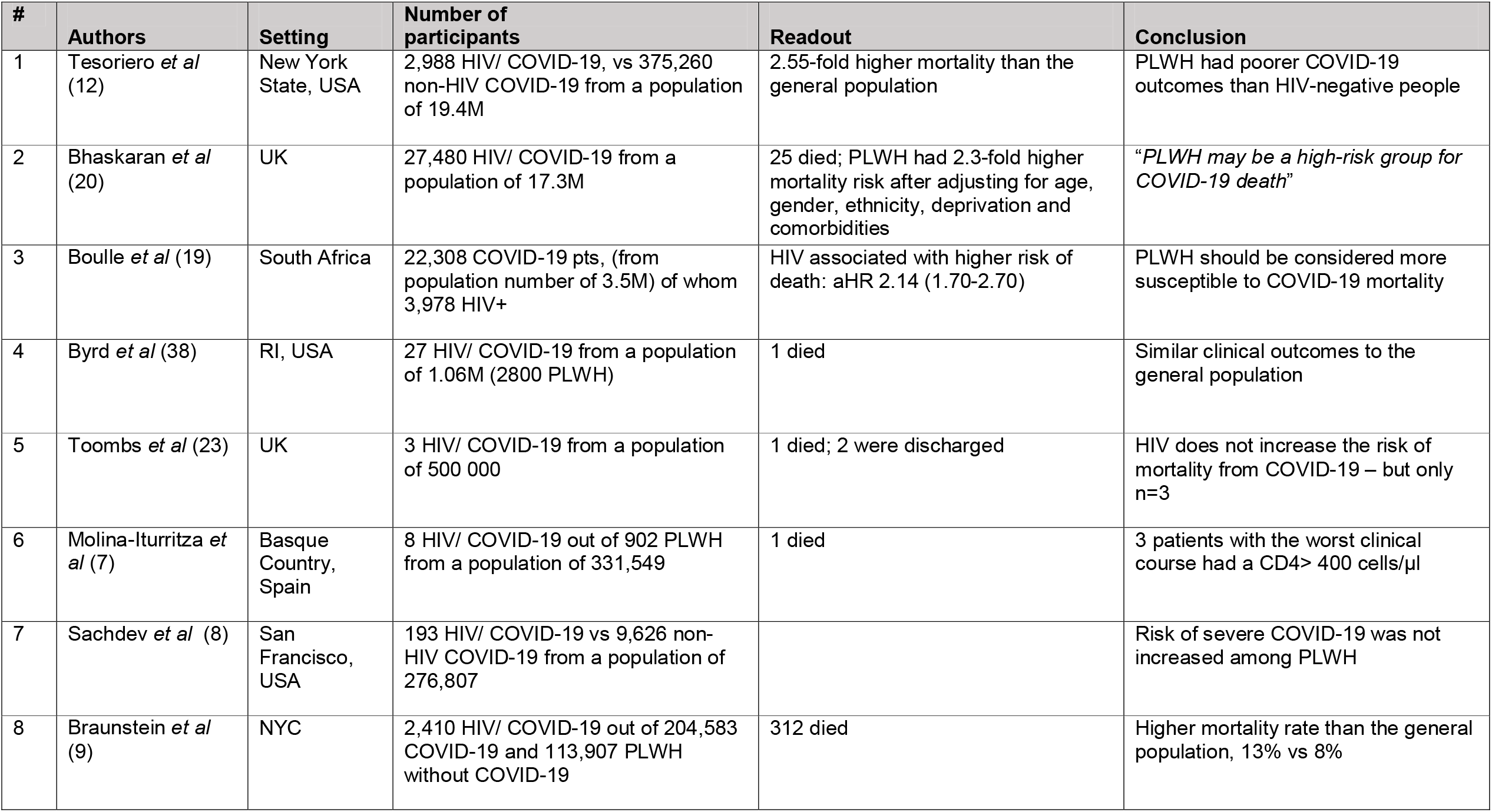
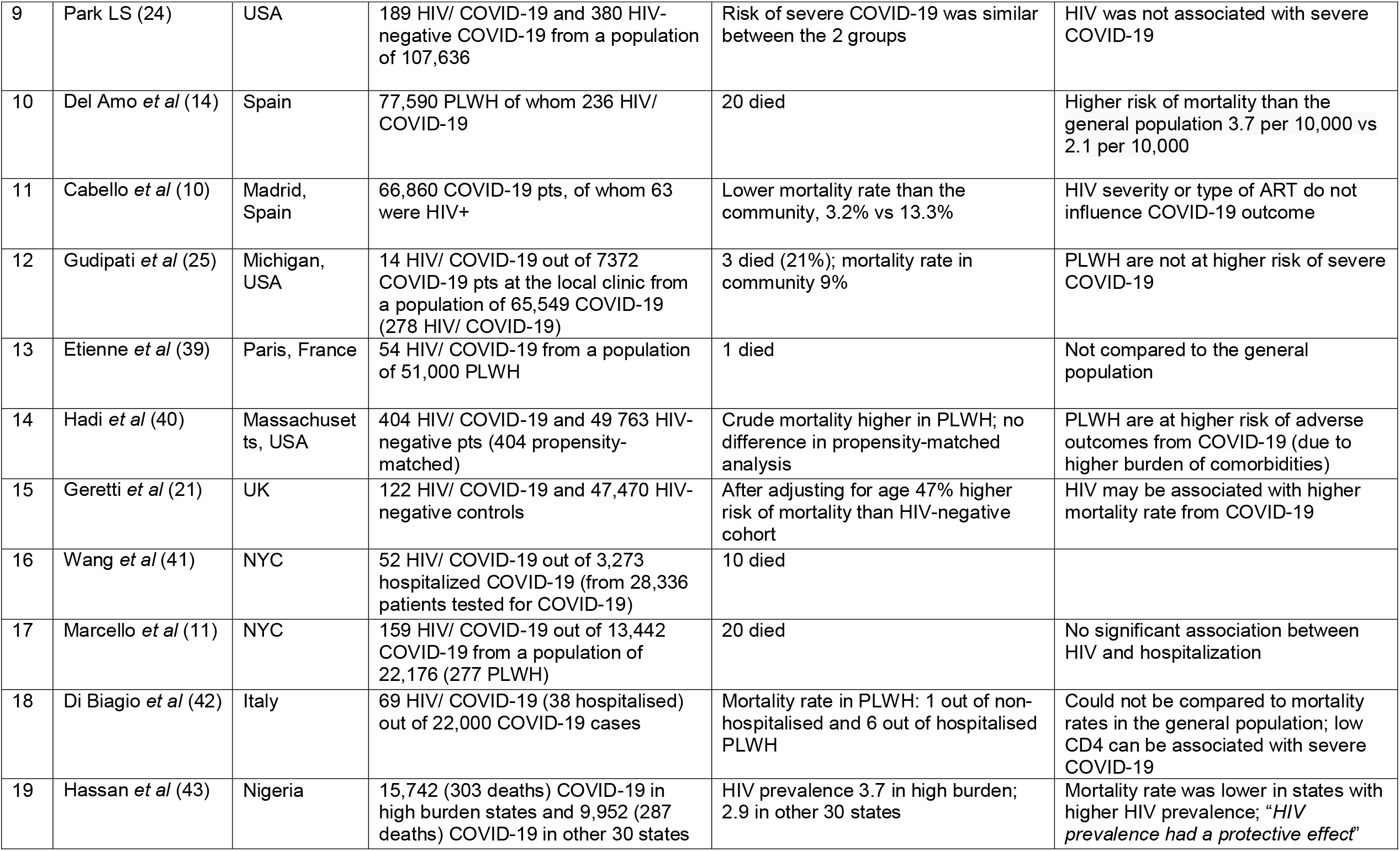
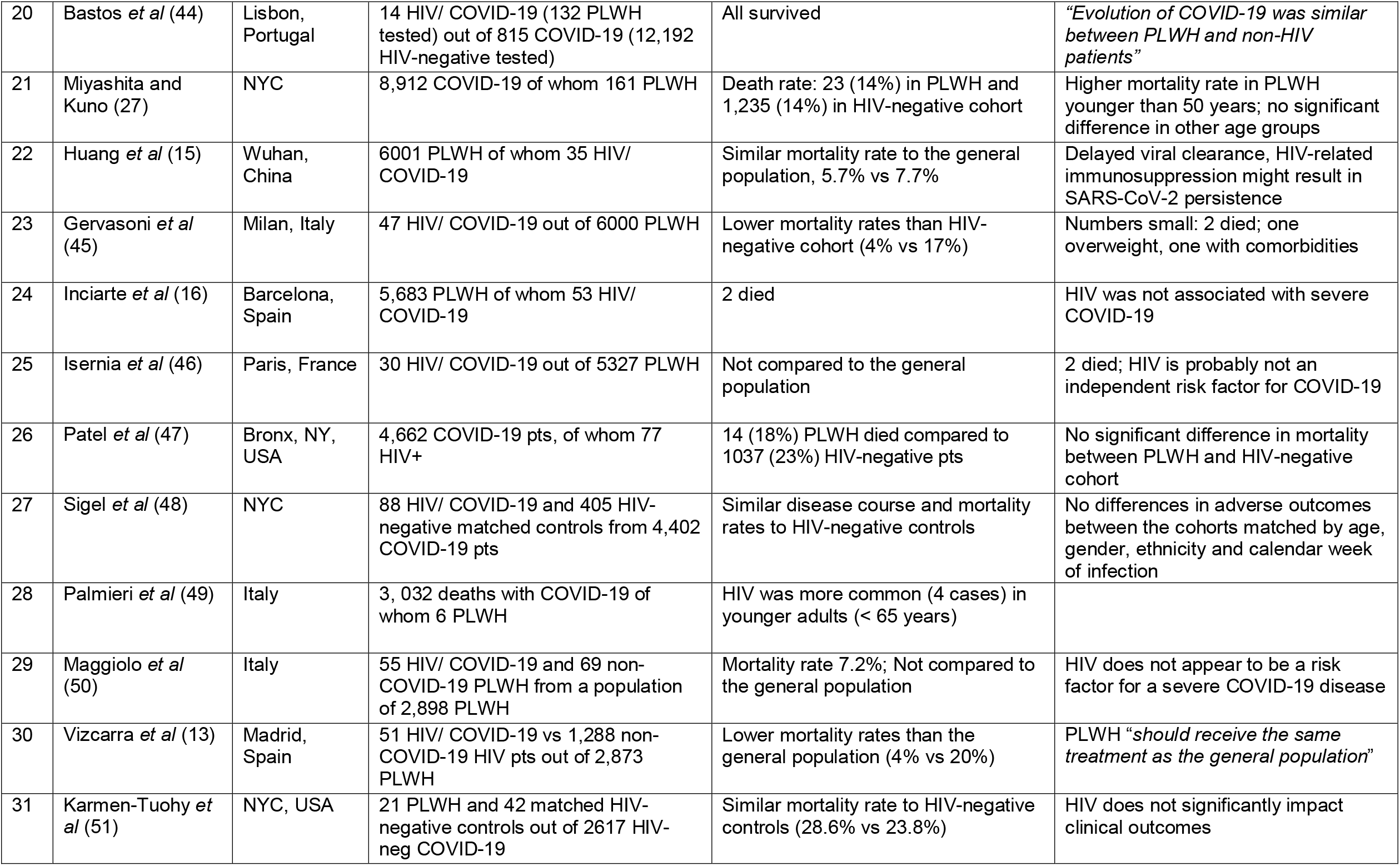
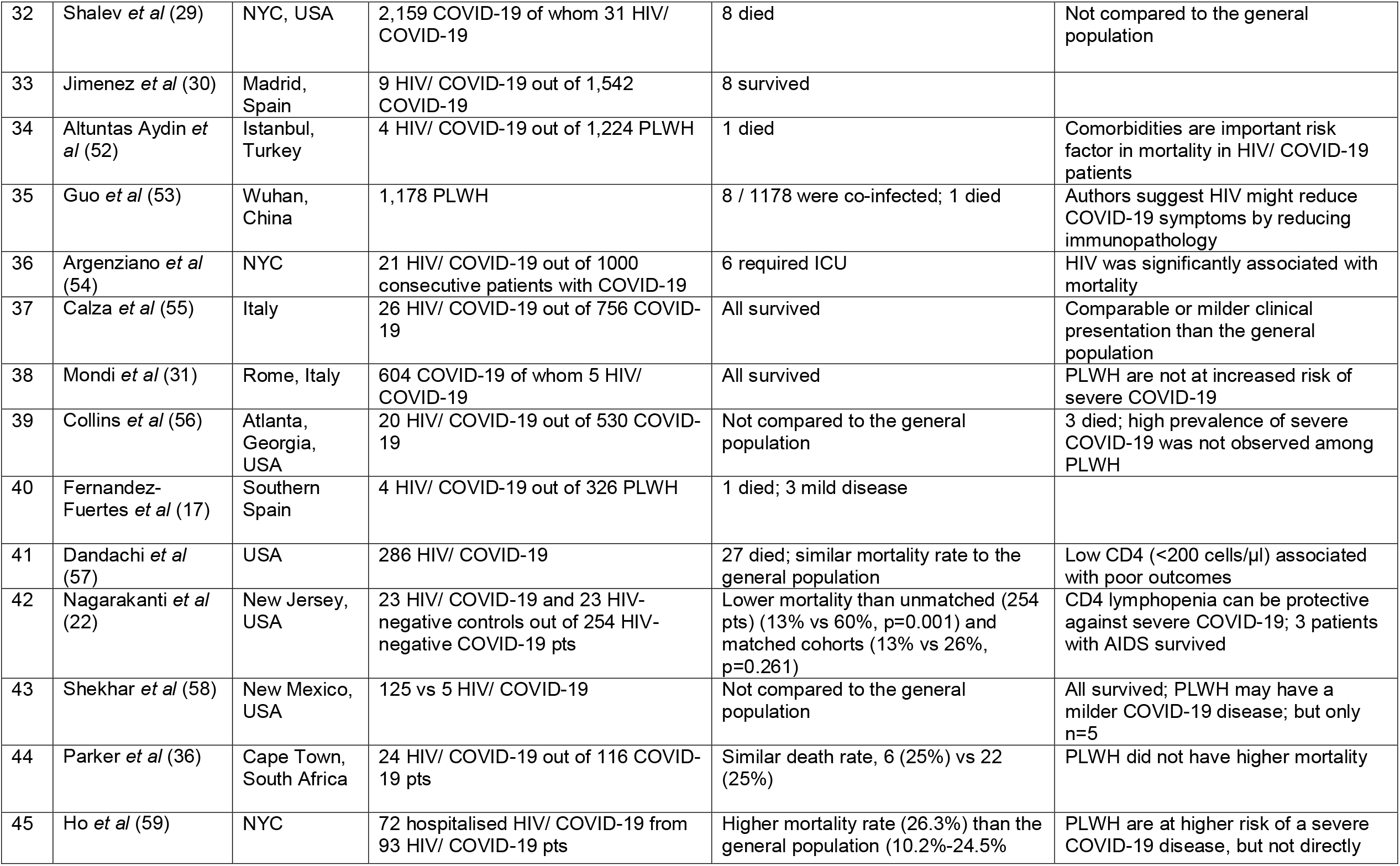
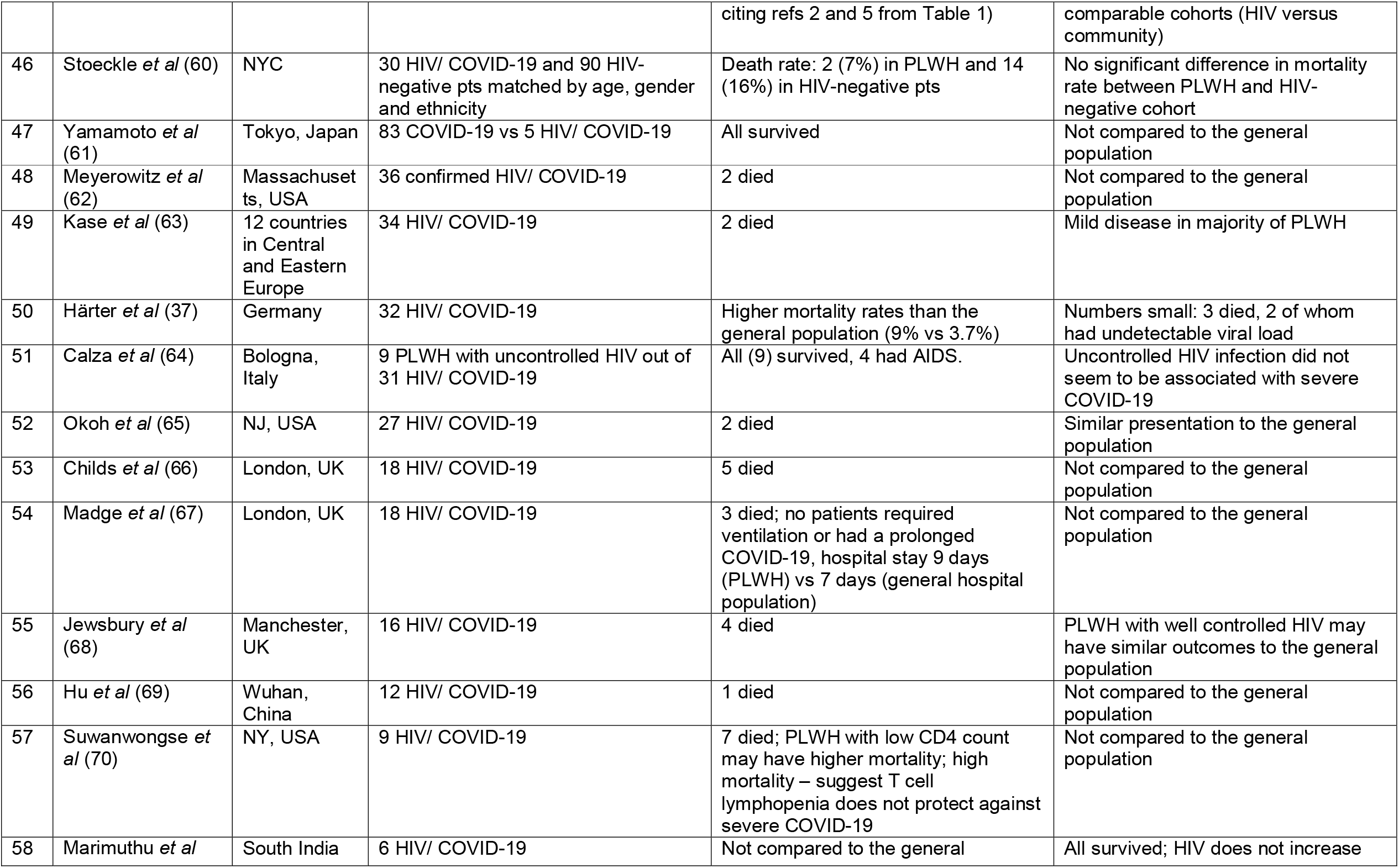
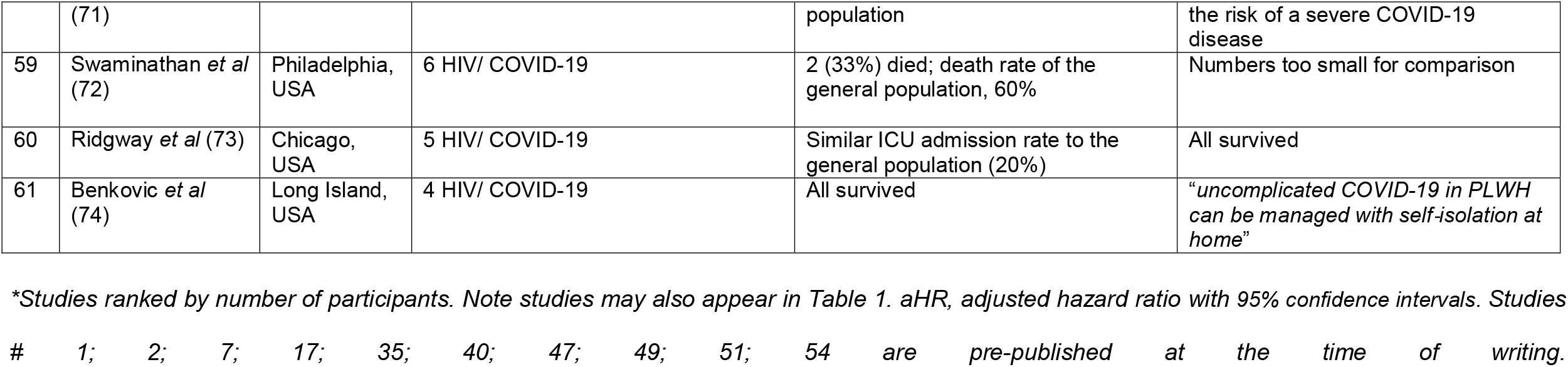
HIV/ COVID-19 co-infection studies with mortality data.

The literature broadly splits into two camps. On one hand, there are a large number of small studies, many of which emerged early in the pandemic. Most cite comparisons of crude mortality rates in PLWH versus the background population; generally, they are not powered to control for other comorbidities. However, in the absence of better data, these papers contributed to a general consensus that HIV does not predispose to more severe disease or higher mortality; as one author put it, there is an *“… under-representation of people living with HIV (PLWH) among severe COVID-19 cases*”. (18) More recently, four studies have been published with much larger numbers of cases that appear to overturn this relatively benign paradigm, the papers by Boulle *et al*, (19) Bhaskaran *et al*, (20) Geretti *et al*, (21) and Tesoriero *et al*, (12) which we will consider in more detail.

Boulle *et al* (19) studied 22,308 public-sector patients with laboratory-confirmed COVID-19 in Western Cape Province, South Africa where the background HIV prevalence is ∼7%. Of these 3,978 (18%) were PLWH. 625 patients died, including 115 PLWH (18%). Although crude mortality rates are almost identical, age at death was lower in PLWH. Hence, when mortality risk was adjusted for risk factors, including age and gender, it became apparent that, in this cohort, HIV doubled COVID-19 mortality rates (aHR 2.14; 95%CI 1.70-2.70), although the authors acknowledge that the risk might be overestimated due to residual confounding. (Current or past TB also increased COVID-19 mortality.) Interestingly, the effect appeared greater in non-hospitalised patients being “*… progressively attenuated when restricting to cases (people with sufficiently severe symptoms to be tested) and hospitalised patients*”. (19)

Another population-based study reviewed primary care data from ∼17M people in the UK, of whom 0.16% were HIV-infected (a relatively low proportion as London, where HIV-positivity rates are highest, was under-represented). (20) Of 14,882 people who died from COVID-19-related causes (according to cause of death records) in the first wave of the pandemic, 25 were HIV-positive (0.17%). The strength of this study is the wealth of co-morbidity data captured. Adjusting just for age and gender gave a hazard ratio of death of 2.9 for PLWH; when all comorbidities were considered the HR was 2.3. Strikingly, the mortality risk from HIV was greater among people of black ethnicity (HR = 3.8). CD4 and viral load data were not available but, in the study setting, ∼94% of PLWH would be expected to be on effective ART.

Considering only hospitalised patients, Geretti *et al* (21) compared 122 HIV/ COVID-19 co-infected patients with 47,470 HIV-negative controls in a large UK-based study (the ISARIC WHO CCP study). PLWH were relatively younger with fewer comorbidities than the HIV-negative cohort. Although the crude cumulative 28-day mortality was similar between PLWH and HIV-negative patients (26.7% vs. 32.1%, p=0.16), after adjusting for age, PLWH had a 47% higher mortality rate. After adjusting for further variables (sex, ethnicity, age, baseline date, indeterminate/ probable acquisition of COVID-19, ten comorbidities, hypoxia/receiving oxygen at presentation) HIV was associated with a 69% higher mortality. The authors could not assess the impact of HIV-related parameters on COVID-19 outcomes as they did not have details of HIV viral load, CD4 count or ART history but they did observe that documented ART was associated with lower mortality.

The Tesoreiro *et al* study, (12) summarised above also reported mortality risk. Interestingly, in their study, the mortality risk once patients were hospitalised was similar to non-HIV infected individuals, but taking increased diagnosis and admission rates into account, the standardized mortality ratio for PLWH was higher than the general population (1.23; 95% CI: 1.13-1.48). As in Geretti *et al*, (21) hospitalized and fatal COVID-19 cases were younger among PLWH.

### Do viral load and CD4 status influence COVID-19 risk?

In many of the studies reviewed, HIV infection is conflated to a single entity. It is clear however that there is a great disparity between someone with suppressed disease (undetectable viral load, high CD4 count) and someone with advanced disease (low CD4 count) and/or an uncontrolled viral load. Many studies have either had insufficient numbers to draw conclusions or been unable to address this issue because of their data collection approach. In terms of data, if we consider CD4 count during the episode as more reflective of COVID-induced lymphopenia than the underlying HIV-related immune deficit, we are left with a relatively small number of studies with CD4 count data preceding the COVID-19 diagnosis. Examples include the Spanish COVID-19 incidence study cited above, (13) which reported a trend linking lower CD4 counts with more severe disease (422 vs 668 cells/uL) but was not significant (p=0.115). In the NYC study, COVID-19 diagnosis rates were higher in those with viral suppression (perhaps reflecting greater engagement with healthcare) as were PLWH with low CD4 counts. In terms of hospitalisation, those with unsuppressed viral loads and low CD4 counts were more likely to be admitted, but CD4 count was not significantly associated with in-hospital death. (12) In the South African study, those in the {VL ≥ 1000 copies/ml (last 15 months) or CD4 <200 cells/µl (last 18 months)} category appeared to have a higher mortality rate, (19) although many patients did not have recent CD4 or viral load measurements, limiting the power of this observation.

## Discussion

In this paper we summarise the extant literature on the interaction of HIV with COVID-19. We used an analytic model separating risk of infection from risk of severe disease. In terms of risk of infection, the definitive answer would come from asymptomatic screening, but HIV status is unlikely to be collected as part of such exercises. Hence, diagnosis rates are the next-best surrogate marker, even though they are profoundly influenced by the occurrence of symptoms or hospitalization. We identified 27 studies to date which address infection risk as a function of HIV status. Most studies assessed HIV prevalence in COVID-19 diagnosed individuals, although some followed COVID-19 incidence in HIV clinic cohorts. (7, 13-17) The consensus that has emerged provides reassurance that PLWH appear no more likely to become infected than their non-infected counterparts; in our model, β_HIV_≈β_0_. The caveat to this conclusion is that PLWH may have been shielding or otherwise reducing their exposure (reducing the factor, α) during the pandemic, in which case, true infection risk may be higher than estimated.

In terms of severity, early studies were reassuring. PLWH did not appear over-represented among those experiencing complications with COVID-19. However, it is now apparent that, when adjusted for other risk factors, even well-controlled HIV infection does appear associated with increased mortality risk. Indeed, much of the disparity between the findings of different studies can be accounted for by the way in which authors have accounted (or not) for co-factors and confounders in their models. Almost all studies that have considered comorbidities have found that the same factors operate within HIV-positive COVID-19-infected cohorts as have been identified in the general population (age, cardiovascular disease, obesity etc), as might be expected. Taking co-factors into account is not possible in small studies, but larger studies allow adjustment which may be crucial for interpretation. So, for example, in Geretti *et al* (21) and Bhaskaran *et al*, (20) unadjusted mortality rates in PLWH are no higher than background mortality rates. However, in both studies, once data are adjusted for age etc, the independent adverse impact of HIV infection is evident. This increased mortality risk with HIV infection manifests itself in different parts of the patient pathway in different settings. Thus in the South African study cited above, the increased mortality risk was most apparent in non-hospitalised individuals; (19) in the NYC study, the big difference was in diagnosis and hospitalisation rates - once admitted mortality was similar to non-HIV-infected individuals; (12) in the UK study, in which hospitalisation was an entry criterion, mortality rates were found to be significantly higher in PLWH after admission. (21)

It does therefore appear reasonable to take HIV status into account in developing policies and practice, but do we treat all PLWH the same? What about the impact of CD4 count and viral load? Here, the data is far less clear-cut. We can postulate with some confidence that the additional risk of HIV infection for mortality seems to apply even to those with well-controlled HIV. Several studies observe an independent effect of HIV infection across all strata; for example Tesoriero *et al* found an increased hospitalisation risk in individuals with CD4≥500 and viral suppression. (12) Furthermore, the two UK studies discussed above were performed in settings where 94% of subjects would be expected to be well-controlled. Whether those with low CD4 counts and/or viraemia are at higher risk remains unclear. Several studies show a relationship between lower CD4 counts and worse outcomes, (12-13,19), but some studies suggest low CD4 counts are protective. (7, 22) Although it might appear counter-intuitive that “immunosuppression” would worsen “immunopathology”, it should be remembered that advanced HIV is characterised by immune dysfunction with heightened levels of immune activation as much as “immunosuppression”.

Recommendations based upon CD4 and/or viral load status thus appear to have a weak evidence-base, but may be justified in a pragmatic risk-stratified model. Current UK guidelines differentiate those with a CD4 count <50 cells/uL for special consideration, (3) but there is little evidence supporting where to make this cut-off. In the NYC study, the authors separated out a cohort with a CD4 count <200 cells/uL or <14% (“Stage 3”) and found they had a higher HIV-attributable mortality risk, (12) but they did not explore lower cut-offs. At the other end of the scale, it does not appear possible to define a “safe” level of CD4 count from the studies published to date; indeed, one can argue from the mortality impact of HIV even in settings where almost everyone is on effective treatment, (20-21) that no such “safe” level exists. One might however pragmatically define a level where recommendations for PLWH match those for the general population. Such stratified advice is likely to vary on a regional basis. Currently, the countries with most COVID-19 cases are relatively low HIV seroprevalence countries. Conversely, countries with high HIV seroprevalence may have limited COVID-19 testing resources and incidence rates may be underestimated. This is why studies such as Boulle *et al* (19) are particularly valuable as they address COVID-19 risks in a high HIV burden setting (as well as making intriguing observations about TB/COVID interactions). Although their conclusions may not be generalisable to other settings, the consistency in message, if not magnitude, of the effect of HIV on COVID-19 mortality across all the larger studies cited is notable.

This overview has attempted to be comprehensive, but we may have missed publications and have not captured those in languages other than English. Of course, in such a rapidly-changing situation new studies are being published every month but this study summarises the current state-of-play and constitutes a comprehensive basis for future literature reviews. We have deliberately chosen not to focus on whether some antiretroviral drugs impact COVID-19 outcomes directly as the data are conflicting and such questions are best addressed in prospective clinical trials. We considered a meta-analysis, but after reviewing the disparate nature of studies with their different data collection strategies, and estimating the number of studies that would need to be excluded, felt that a comprehensive descriptive analysis was more helpful.

Understanding the impact of HIV on COVID-19 is important in order to inform guidance and policy. “Shielding” has costs in terms of emotional and psychological wellbeing, finances and quality of life. If PLWH are to be asked to take extra precautions, this guidance needs to be based on good data. Additionally, as vaccines become available, prioritisation for vaccination is likely, in which case we need data to establish where PLWH stand in any risk-stratification. Despite an extensive body of literature, more research is needed to clarify risk in different settings and particularly to clarify how guidance should be shaped by measures such as CD4 count and viral load, but the new data from large studies is consistent: even well-controlled HIV appears associated with an increased COVID-19 mortality risk.

## Data Availability

Data available within the article or its supplementary materials.

## Acknowledgements

We acknowledge the help of Rebecca Marchant, Lisa Hamzah and Padmini Parthasarathi in evaluating local rates of COVID-19 among PLWH.

